# Inflammatory markers in Covid-19 Patients: a systematic review and meta-analysis

**DOI:** 10.1101/2020.04.29.20084863

**Authors:** Golnaz Vaseghi, Marjan Mansourian, Raheleh Karimi, Kiyan Heshmat-Ghahdarijani, Paria Rouhi, Mahfam Shariati, Shaghayegh Haghjoo Javanmard

## Abstract

**Introduction:** Diagnosis of COVID-19 is based on clinical manifestation, history of exposure, positive findings on chest CT and laboratory tests. It has been shown that inflammation plays a role in pathogenesis of COVID-19.

**Method:** We used the necessary transformations to convert the median and IQR to mean and SD Random-effect model using Der Simonian, and Laird methods was used if heterogeneity between studies was significant, the homogeneity among studies was assessed with I^2^ Statistic, values above 50%, and for the chi-square test, P-values <0.1 was supposed statistically significant

**Results:** Twelve studies were included in the analysis that all of which were conducted in China in the year 2020. The result of combining 12 articles with 772 participants showed that the pooled estimate of the mean of lymphocyte with 95% CI was (Mean: 1.01; 95% CI (0.76-1.26); p-value<0.001). About WBC the pooled result of 9 studies with 402 participants was (Mean: 5.11; 95% CI (3.90-6.32); p-value<0.001) Also the pooled mean estimate of 9 studies with 513 patients for the ratio of Neutrophil/lymphocyte was (Mean: 3.62; 95% CI (1.48-5.77); p-value=0.001). The pooled mean from the combination of 7 studies with 521 patients on CRP was (Mean: 28.75; 95% CI (8.04-49.46).

**Conclusion:** Inflammatory Markers increase in patients with Covid-19, which can be a good indicator to find patients.

## Introduction

A novel coronavirus was identified as the causative pathogen of pneumonia in Wuhan City, Hubei province in China in December 2019 (1). It was subsequently named Severe Acute Respiratory Syndrome Coronavirus 2 (SARS-CoV-2). The disease was named COVID-19 by the World Health Organization (WHO) (2). On 30th January 2020, the WHO declared the Chinese outbreak of COVID-19 to be a Public Health Emergency of International Concern posing a high risk to countries with vulnerable health systems (3). To date many countries in the world in different continents was involved in COVID-19 and its complications and it became a pandemic disease.

Most patients with COVID-19 have developed mild symptoms such as dry cough, sore throat, fatigue and fever. The majority of cases have spontaneously resolved. However, some patients developed fatal complications such as organ failure, septic shock and Acute Respiratory Distress Syndrome (ARDS) and finally died (4).

Diagnosis of COVID-19 is based on clinical manifestation, history of exposure, positive findings on chest CT and laboratory tests (5). It has been shown that inflammation plays a role in pathogenesis of COVID-19 (6). In these patients release of pro-inflammatory cytokines followed by inflammasome activation produce lung injury (7).

Systemic inflammation changes the features of circulating blood cells have been suggested to be biomarkers for assessment of inflammatory activity. Neutrophilia with lymphopenia is a response of the innate immune system to systemic inflammation. The neutrophil-to-lymphocyte ratio (NLR) is another inflammatory marker, which is the proportion of absolute neutrophil count to lymphocytes on routine complete blood count (CBC) tests. An elevated NLR is associated with the prognosis of systemic inflammatory diseases, especially infectious diseases (8).

CRP as the most representative of acute phase reactants, is elevated in many respiratory viral infections (9). CRP testing is used at the forefront for evaluation of infection and is important for COVID-19 management (10).

In this meta-analysis, we decided to evaluate the White blood cell and lymphocyte count, NLR and CRP titer in COVID-19 to use these findings in the diagnosis of the COVID-19 more rapidly and accurately.

## Method

### Literature search and selection criteria

We used Systematic Reviews and Meta-Analyses Statement for Scoping Reviews” (PRISMA-ScR) (11).

Pubmed, EMBASE and Scopus, were searched for eligible publications, till March 20, 2020. The search strategy was ((((((coronavirus[MeSH Terms]) OR coronavirus infections[MeSH Terms]) OR “betacoronavirus”[MeSH Terms]) OR “betacoronavirus 1”[MeSH Terms]) OR (Coronaviruses OR “Coronavirus Infection” OR “COVID-19” OR “Coronavirus Infection Disease 2019” OR “2019 Novel Coronavirus Infection” OR “2019 nCoV Infection” OR “2019 nCoV Infection” OR “2019-nCoV Infections” OR Betacoronavirus* OR “Novel Coronavirus Pneumonia” OR “2019 novel coronavirus” OR “coronavirus disease 2019” OR “nCoV” OR covid* OR “bat coronavirus”)). We included all case control, editorial and epidemiological studies. Titles and abstracts of all searches were screened independently by two investigators. Articles deemed potentially eligible were retrieved for full-text review. Non-English publications were translated by a native/fluent speaker.

### Statistical methods

Two measures of association were used in studies; mean (SD) and median (IQR). We used the necessary transformations to convert the median and IQR to mean and SD (12). Therefore, the pooled estimation is reported based on mean and a corresponding 95% CI. Random-effect model using Der Simonian and Laird methods was used if heterogeneity between studies was significant, to Summary the results, otherwise a model with fixed effect was calculated (13). The homogeneity among studies was assessed with I^2^ Statistic, values above 50%, and for the chi-square test, P-values <0.1 was supposed statistically significant (14). Publication bias was evaluated with Begg’s rank correlation tests and Egger’s linear regression tests (15). If the test for Publication bias become significant, trim and fill methods were used to modify the results. All analyses were performed using STATA version 14.

## Result

Figure. 1 shows the search strategy result. Twelve studies were included in the analysis that all of which were conducted in China in the year 2020. The age of patients varied from 40 to 75 years. Summary information of each article is presented in Table. 1, As shown in Table.2, The result of combining 12 articles with 772 participants showed that the pooled estimate of the mean of lymphocyte with 95% CI was (Mean:1.01; 95% CI (0.76-1.26); p-value<0.001, Figure. 2). About WBC the pooled result of 9 studies with 402 participants was (Mean: 5.11; 95% CI (3.90-6.32); p-value<0.001) (the forest plot is presented in Figure. 3). Also the pooled mean estimate of 9 studies with 513 patients for the ratio of Neutrophil/lymphocyte was (Mean: 3.62; 95% CI (1.48-5.77); p-value=0.001, Figure. 4). The pooled mean from the combination of 7 studies with 521 patients on CRP was (Mean: 28.75; 95% CI (8.04-49.46); p-value<0.001, Figure. 5. The heterogeneity test was not significant for any of the outcomes (p-value>0.05), so all results were based on a fixed model. Also the publication bias except for the WBC (p-value=0.026), for others were not significant. The adjusted result of Trim and fill test to modify the result of WBC, was (Mean: 4.97; 95% CI (3.81-6.13)).

**Figure. 1:**
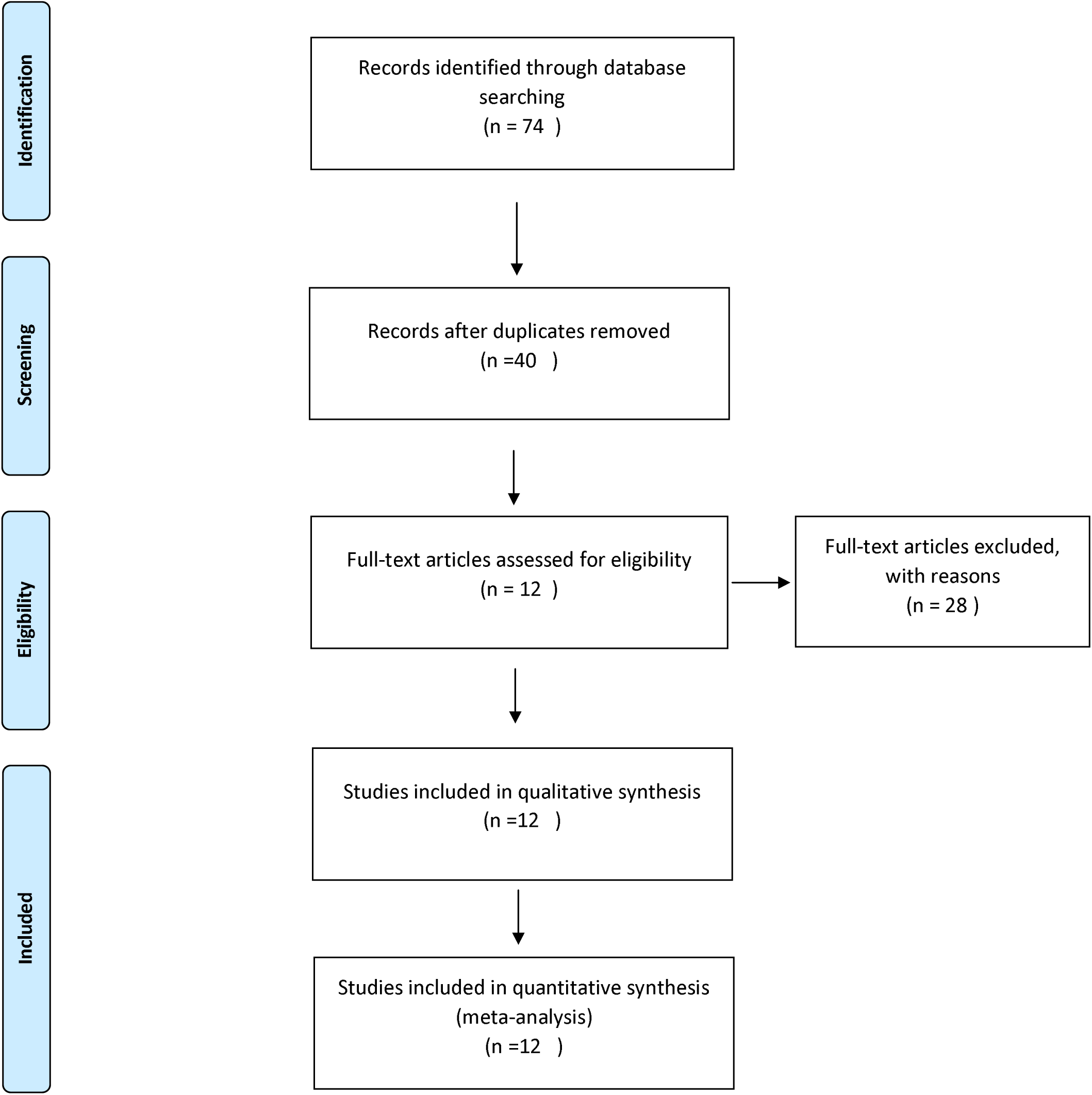
Flow diagram

**Figure. 2:**
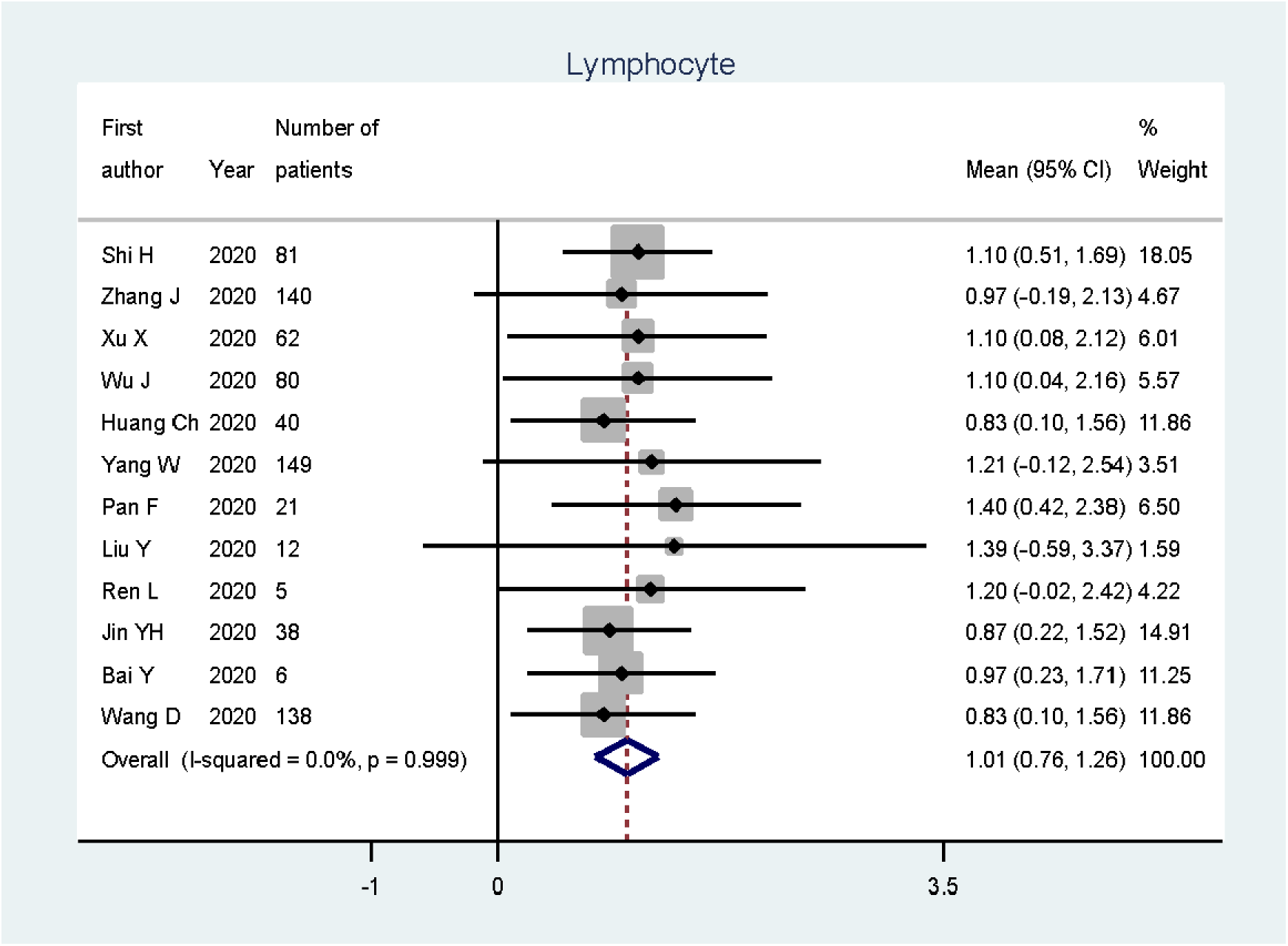
Forest plot of overall mean of Lymphocyte in COVID-19 patients

**Figure. 3:**
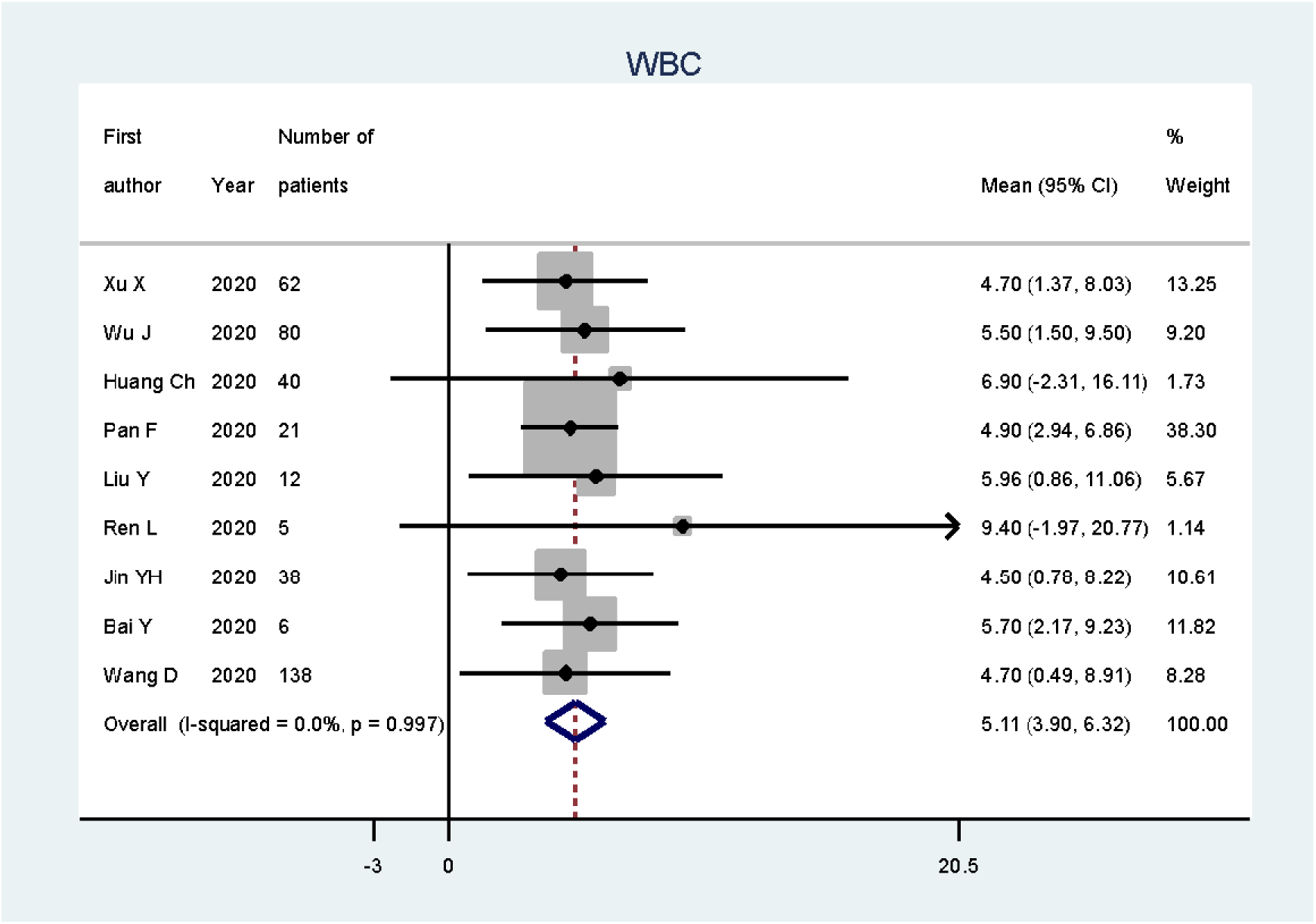
Forest plot of overall mean of WBC in COVID-19 patients

**Figure. 4:**
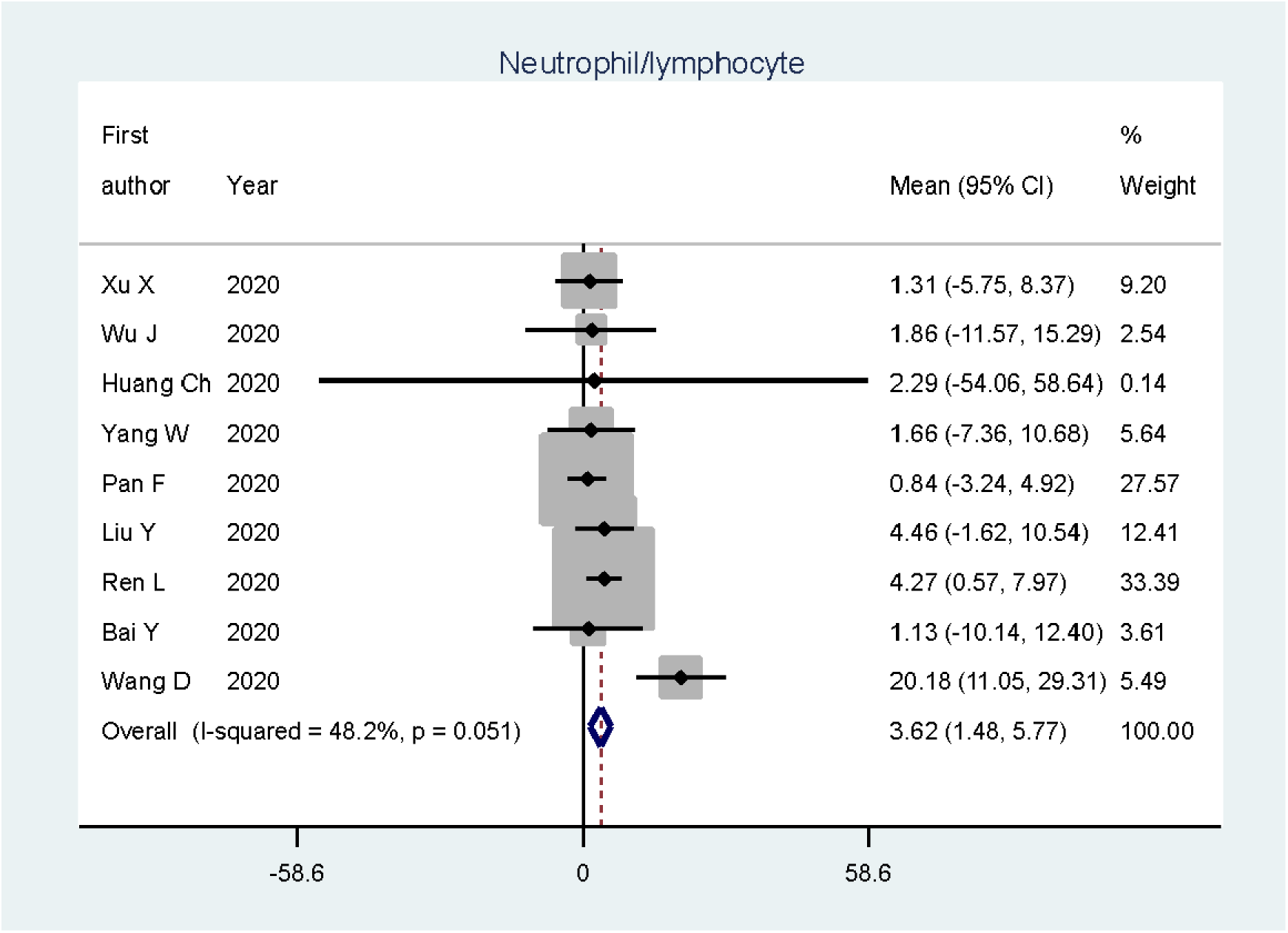
Forest plot of overall mean of Neutrophil/lymphocyte in COVID-19 patients

**Figure. 5:**
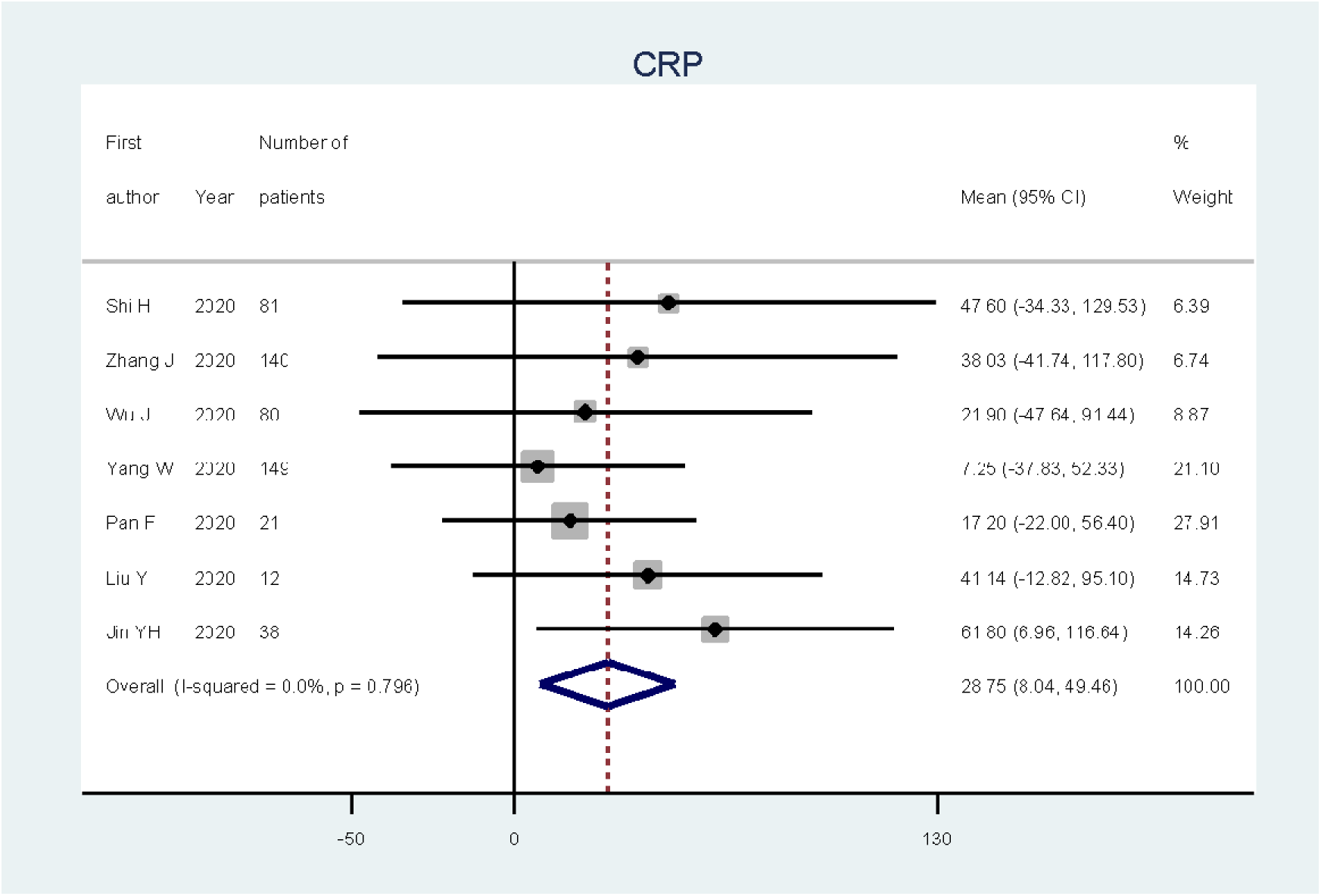
Forest plot of overall mean of CRP in COVID-19 patients

**Table 1:**
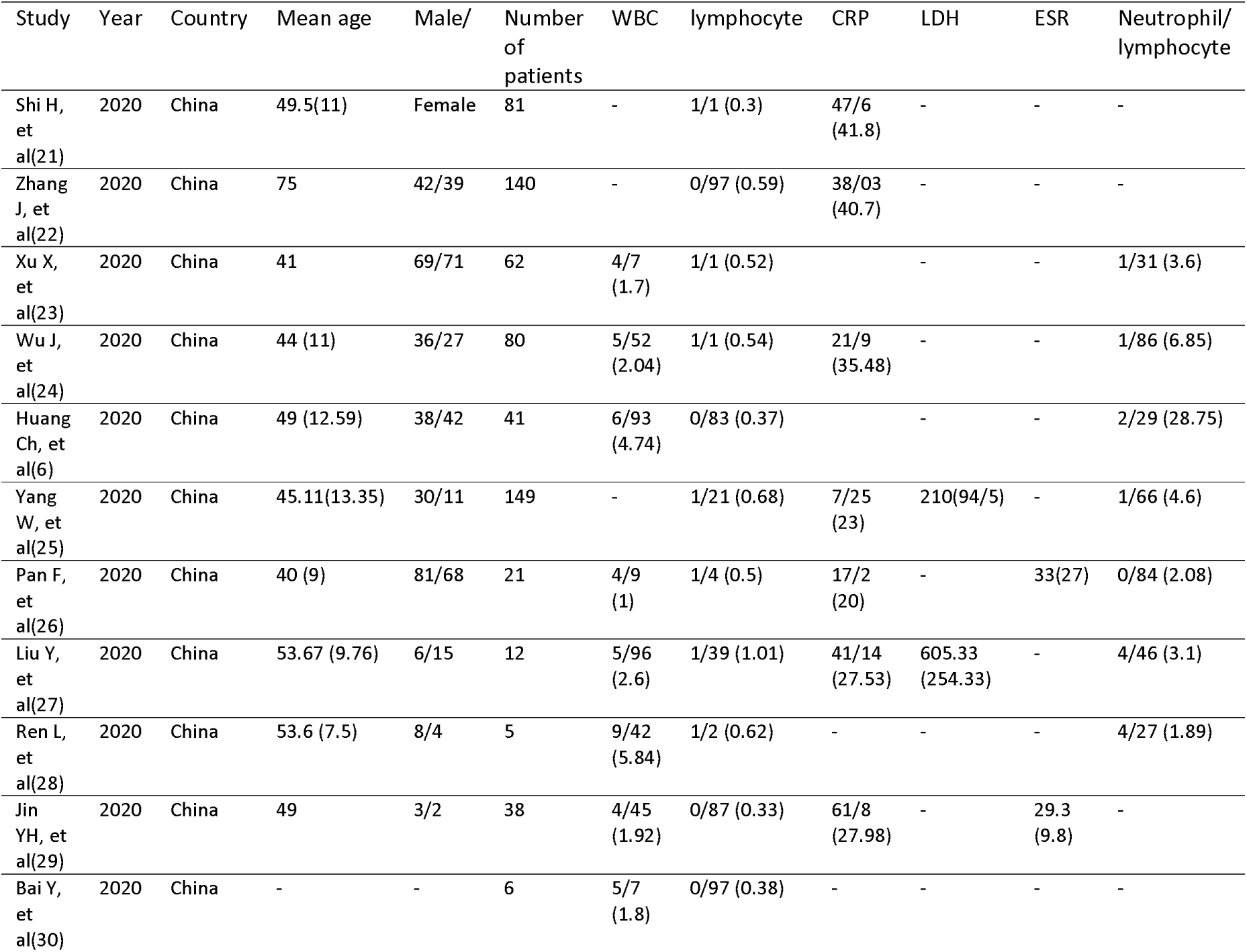
studies information

| Table1: studies information |  |  |  |  |  |  |  |  |  |  |  |
| --- | --- | --- | --- | --- | --- | --- | --- | --- | --- | --- | --- |
| Study | Year | Country | Mean age | Male/<br>Female | Number<br>of<br>patients | WBC | lymphocyte | CRP | LDH | ESR | Neutrophil/<br>lymphocyte |
| Shi H, et al(21) | 2020 | China | 49.5(11) | Female | 81 | - | 1/1 (0.3) | 47/6 (41.8) | - | - | - |
| Zhang J, et al(22) | 2020 | China | 75 | 42/39 | 140 | - | 0/97 (0.59) | 38/03 (40.7) | - | - | - |
| Xu X, et al(23) | 2020 | China | 41 | 69/71 | 62 | 4/7 (1.7) | 1/1 (0.52) |  | - | - | 1/31 (3.6) |
| Wu J, et al(24) | 2020 | China | 44 (11) | 36/27 | 80 | 5/52 (2.04) | 1/1 (0.54) | 21/9 (35.48) | - | - | 1/86 (6.85) |
| Huang Ch, et al(6) | 2020 | China | 49 (12.59) | 38/42 | 41 | 6/93 (4.74) | 0/83 (0.37) |  | - | - | 2/29 (28.75) |
| Yang W, et al(25) | 2020 | China | 45.11(13.35) | 30/11 | 149 | - | 1/21 (0.68) | 7/25 (23) | 210(94/5) | - | 1/66 (4.6) |
| Pan F, et al(26) | 2020 | China | 40 (9) | 81/68 | 21 | 4/9 (1) | 1/4 (0.5) | 17/2 (20) | - | 33(27) | 0/84 (2.08) |
| Liu Y, et al(27) | 2020 | China | 53.67 (9.76) | 6/15 | 12 | 5/96 (2.6) | 1/39 (1.01) | 41/14 (27.53) | 605.33 (254.33) | - | 4/46 (3.1) |
| Ren L, et al(28) | 2020 | China | 53.6 (7.5) | 8/4 | 5 | 9/42 (5.84) | 1/2 (0.62) | - | - | - | 4/27 (1.89) |
| Jin YH, et al(29) | 2020 | China | 49 | 3/2 | 38 | 4/45 (1.92) | 0/87 (0.33) | 61/8 (27.98) | - | 29.3 (9.8) | - |
| Bai Y, et al(30) | 2020 | China | - | - | 6 | 5/7 (1.8) | 0/97 (0.38) | - | - | - | - |

**Table. 2:** summery of the results

| Table 2: summery of the results |  |  |  |  |  |  |  |  |
| --- | --- | --- | --- | --- | --- | --- | --- | --- |
|  | No of<br>Study | Number<br>of<br>subject | Mean [95%<br>CI] | P-value for<br>test (ES=0) | I<sup>2</sup><br>(%) | P-value for<br>heterogeneity<br>test | P-value<br>for egger<br>test | Trim and fill<br>results<br>(%95CI) |
| **WBC** | 9 | 402 | 5.11 (3.90-6.32) | < 0.0001 | 0.0 | 0.997 | 0.026 | 4.97 (3.81-6.13) |
| **lymphocyte** | 12 | 772 | 1.01 (0.76-1.26) | < 0.0001 | 0.0 | 0.999 | 0.067 | - |
| **Neutrophil/**<br>**lymphocyte** | 9 | 513 | 3.62 (1.48-5.77) | 0.001 | 48.2 | 0.051 | 0.650 | - |
| **CRP** | 7 | 521 | 28.75 (8.04-49.46) | 0.007 | 0.0 | 0.796 | 0.240 | - |

## Discussion

Several studies have suggested that increased amounts of cytokines in serum are associated with pulmonary damage in SARS and MERS-CoV infection (16, 17).

In this meta-analysis, we studied the inflammatory markers of CBC and lymphocyte upset in patients with COVID-19. We included all epidemiological and case report studies. The patients age was between 40 to 75 years.

The blood level of WBC was normal with reduced amount of lymphocyte, as expected (18). In terms of laboratory tests, we noted that most of infected patients presented lymphopenia and low counts of CD3+ cells and CD4+ cells have been observed in COVID-19 cases (19). NLR is used as a marker of subclinical inflammation, especially in severe pneumonia (20). Here we showed that the pool NLR of COVID-19 is 3.62 which is upper than physiological range.

An increase of the NLR is common among patients with COVID-19, and especially in the severe cases, COVID-19 damages lymphocytes and the immune system. It has been suggested that NLR is the most significant factor affecting the severe illness incidence, and it has significant predictive value (19).

Based on the previous studies, Patients with 2019-nCoV pneumonia with higher NLR and age would face a worse outcome which helps their risk stratification. If there are large-scale cases, the risk stratification and management will help alleviate the shortage of medical resources and reduce the mortality of critical patients.

In conclusion, in this study, we found that patients with COVID-19 have higher CRP and NLR but low lymphocyte and normal WBC, which can help in diagnosis of these patients.

## Data Availability

All the data is available in the full text article

## Acknowledgements

Authors would like to thank all health care workers who fight COVID-19.

